# Mapping the Diagnostic Space of Auditory Brainstem Responses: An Interpretable Framework for Classification

**DOI:** 10.64898/2026.09.14.26363001

**Authors:** Letizia Clementi, Daniele Cazzato, Elisa Visani, Marie Carolina Nicolis di Robilant, Annamaria Gallone, Virginia Iacobelli, Paola Lanteri, Davide Rossi Sebastiano

## Abstract

**Background:** Auditory Brainstem Responses (ABRs) are objective and highly standardized evoked potentials, but their clinical interpretation still largely depends on expert visual assessment. Most automated ABR approaches focus on signal detection, threshold estimation, or waveform recognition, whereas the diagnostic reasoning linking ABR features to clinically meaningful categories remains insufficiently formalized.

**Methods:** We developed a theoretical, rule-based framework for ABR classification. Nine clinically relevant descriptors were defined: amplitudes of waves I, III, and V; latencies of waves I, III, and V; the I/V amplitude ratio; and the I-V and III-V interpeak intervals. The complete theoretical combinatorial space was generated and progressively reduced using rules that accounted for non-applicable descriptors, algebraic consistency, and neurobiological plausibility. Two expert raters independently labeled valid configurations and compared with language-model-assisted and logic-based rule systems. Agreement was assessed using percentage concordance and Cohen’s κ. Descriptor importance was explored using Random Forest analysis.

**Results:** The initial space of 5,832 possible ABR configurations was reduced to 343 valid configurations. Expert raters showed high agreement, with 84.55% concordance and Cohen’s κ = 0.620. Agreement between expert classifications and language-model-assisted rules was lower, whereas logic-based rules showed higher agreement with both raters and were retained as the final proposed rule set. The diagnostic space was highly asymmetric, with NORM, PPSHA, and PPC occupying narrow regions, and PB and MIXED representing broader diagnostic domains. Wave I amplitude was the most influential descriptor across classification agents.

**Conclusions:** This work provides an interpretable framework for ABR classification by transforming a broad theoretical combinatorial space into a constrained diagnostic domain. This proof-of-concept study supports the feasibility of modelling ABR interpretation as a structured diagnostic space grounded in auditory neurobiology, providing a reproducible foundation for future clinical validation and semi-automated decision-support tools.

**Highlights:**

- Auditory Brainstem Response (ABR) interpretation can be formalized as a constrained diagnostic space.
- A theoretical space of 5,832 ABR configurations was reduced to 343 valid patterns.
- Logic-based rules provided an interpretable bridge between ABR descriptors and diagnostic classification.

## Introduction

Evoked potentials (EPs) represent an ideal testbed for the development of automated, interpretable neurophysiological decision-support systems. EPs are elicited by controlled stimuli, time-locked to the event of interest, and usually interpreted through the analysis of a limited set of reproducible waveform components, which are characterized by a standardized set of features including latencies, amplitudes, morphology, and interpeak intervals. This combination of standardization, anatomical interpretability, and clinically meaningful quantitative descriptors makes EPs particularly suitable for computational modelling, formalization, and interpretation supported by machine-learning.

Within this wider domain, Auditory Brainstem Responses (ABRs) offer one of the most favorable models for developing semi-automated neurophysiological classification. They are highly reproducible, have well-established clinical use, and exhibit relatively constrained waveform architecture. Previous studies have already shown that several properties of ABR analysis can be automated, including threshold detection, response identification, waveform recognition, and extraction of relevant signal features through deep-learning (Bogaerts et al., 2009; Suthakar and Liberman, 2019; McKearney and MacKinnon, 2019; Chen et al., 2021; Thalmeier et al., 2022; Liang et al., 2024; Erra et al., 2026). Accordingly, ABRs may act not only as targets for automation, but also as proof-of-concept models for a transition toward interpretable artificial intelligence (AI) in clinical neurophysiology.

ABRs are short-latency EPs generated by the synchronous activation of the auditory nerve and brainstem auditory pathways in response to acoustic stimulation. Since they can be recorded non-invasively and do not require active patient cooperation, ABRs are a widely used objective tool for hearing assessment and for evaluating the functional integrity of the auditory pathway, particularly in newborns, infants, difficult to test patients, and experimental models (Paulraj et al., 2015; Suthakar and Liberman, 2019; Erra et al., 2026). The most relevant components are waves I, III, and V, along with their absolute latencies and amplitudes, amplitude ratios, and interpeak intervals, which provide information on peripheral auditory function and brainstem conduction (Chhajed et al., 2022).

Despite their relatively simple and standardized acquisition, ABR interpretation remains substantially dependent on expert visual assessment. Neurophysiologists must identify wave morphology, estimate latencies and amplitudes, assess interpeak intervals, and then merge these features into a coherent diagnostic interpretation. This process is vulnerable to inter-rater variability, differences in training and experience, signal quality, and local reporting habits (Manta et al., 2022). Several studies have emphasized that visual ABR threshold estimation and waveform interpretation may be subjective, time-consuming, and prone to reader bias (Bogaerts et al., 2009; Suthakar and Liberman, 2019; Thalmeier et al., 2022; McKearney et al., 2022). In this context, a paradox emerges: ABRs are often considered objective neurophysiological tests, however, their ultimate clinical meaning is still largely mediated by subjective expert interpretation.

In recent years, several computational approaches have been proposed to automate different aspects of ABR analysis. Both early and more recent methodological works have focused primarily on objective threshold detection, classification of response presence or absence, and waveform recognition. Automated threshold detection methods have been developed to reduce the variability of visual threshold estimation (Bogaerts et al., 2009; Suthakar and Liberman, 2019; Tanaka et al., 2024; Thalmeier et al., 2022).

Machine-learning and deep-learning models exhibited promising performance in classifying ABR waveforms, as well, detecting responses, estimating thresholds, and discriminating specific waves (Chen et al., 2021; McKearney and MacKinnon, 2019; McKearney et al., 2022; Liang et al., 2024). More recently, convolutional neural network approaches have extended this line of research toward hearing-loss classification from ABR-derived data (Ma et al., 2024), while open-source deep-learning platforms such as ABRA support automated extraction of peak amplitude, latency, and threshold estimates, improving reproducibility and decreasing manual workload (Erra et al., 2026).

However, most present methods remain focused on signal detection, peak recognition, hearing threshold estimation, or coarse normal/pathological classification. These are important technical achievements, but they do not fully reproduce the diagnostic reasoning performed by clinical neurophysiologists. In routine practice, ABR interpretation is not limited to determining whether a response is present or a threshold is abnormal; it also entails mapping combinations of neurophysiological abnormalities onto plausible anatomical lesion sites and pathophysiological processes. Therefore, this clinically oriented, interpretable mapping remains insufficiently formalized in the current computational literature.

The purpose of this work is to address this gap by developing an interpretable and neurobiologically grounded framework for ABR classification. We formalized ABR interpretation as a diagnostic space based on clinically meaningful descriptors, including amplitudes, latencies, amplitude ratios, and interpeak intervals. Each combination of these descriptors represents a possible functional state of the auditory pathway; impossible, inconsistent, or redundant combinations are then excluded, leaving a constrained diagnostic space consistent with auditory neurobiology (Buran et al., 2020).

This approach follows the principle that “there is nothing more practical than a good theory” (Lewin, K., 1951) and, in the context of clinical neurophysiology, a good theory is not an abstract exercise yet a necessary basis to develop automated interpretation transparent, testable, and clinically meaningful, oppositely from black-box machine-learning models, by incorporating domain knowledge into the classifier’s structure (Combi et al., 2022). By combining computational formalization with neurophysiological plausibility, our model is a neuro-symbolic framework in which theoretical diagnostic outputs are generated by transparent logical rules rather than by uninterpretable statistical associations alone.

In this context, interpretability is particularly relevant because it allows the clinical implementation of AI. Across medicine, it is increasingly expected that a support decision system based on AI will not only achieve higher accuracy but will also integrate with medical workflows, support clinical decision-making, and preserve interpretability for clinicians (Susanto et al., 2023; Elhaddad and Hamam, 2024). Similar considerations have emerged in clinical neurophysiology, where automated EEG interpretation has shown that AI can approximate expert-level performance, while likely enhancing consistency and access to specialized-level interpretation to young professionals (Tveit et al., 2023). For ABR analysis, an interpretable rule-based framework may therefore act as a bridge between expert physiological reasoning and future data-driven automation (Tveit et al., 2023).

The present study aimed to develop a theoretical, rule-based diagnostic framework for ABR interpretation. Specifically, we defined a set of clinically relevant ABR descriptors, generated the complete theoretical combinatorial space, applied logical and neurobiological pruning criteria, and assigned the remaining valid configurations to interpretable diagnostic categories. We provide a set of classification criteria, extracted through the application of a Large-Language Model (LLM), and through logical reasoning. This proof-of-concept work provides the logical pillar for future validation of diagnostic impact on clinical ABR recordings and for the development of semi-automated decision-support tools.

## METHODS

### Study design and methodological rationale

The study was conceived as a theoretical, rule-based modelling effort to define an interpretable diagnostic space for ABR. Consistent with the conceptual framework outlined in the Introduction, ABR interpretation was formalized as a constrained multidimensional space in which clinically meaningful neurophysiological descriptors serve as coordinates. The objective of this work was not to train a black-box classifier on clinical waveforms, but to construct a neurobiologically plausible and computationally reproducible map linking ABR feature configurations to diagnostic categories. We did not analyze clinical recordings, rather, all possible configurations of predefined ABR descriptors were generated, filtered, annotated, and translated into formal classification rules.

The methodological workflow comprised the following steps: definition of the ABR descriptor set; generation of the complete theoretical combinatorial space; removal of algebraically inconsistent, neurobiologically implausible, or redundant configurations; expert-based diagnostic annotation of the remaining valid combinations; and extraction of interpretable classification rules. Finally, we characterized the extracted rules. This pipeline was designed to provide the logical foundation for subsequent validation on real-world ABR recordings and for future development of semi-automated decision-support tools. The study design is summarized in the graphical abstract.

### ABR descriptor set

The model focused on waves I, III, and V, which represent the most stable and clinically informative components of the early auditory response (Young et al., 2023). These waves were selected because they capture essential neurophysiological information required to infer peripheral and brainstem conduction (Eggermont JJ, 2019). Consequently, the descriptor set was deliberately restricted to variables commonly considered in routine clinical interpretation to maximize interpretability and translational relevance (Chen et al., 2021).

Nine electrophysiological descriptors were included: amplitudes of waves I, III, and V; absolute latencies of waves I, III, and V; the amplitude ratio between waves I and V; and the interpeak intervals between waves I and V and between waves III and V (Young et al., 2023). These descriptors were encoded as I, III, V, LI, LIII, LV, I/V, IB, and IT, respectively. IB was defined as the III-V interval and was intended to represent conduction through the brainstem, at the pons and midbrain level, of the auditory pathway. IT was defined as the total I-V interval and was intended to represent overall auditory brainstem conduction time (Young et al., 2023).

Each descriptor was treated as a categorical variable reflecting its physiological state. All descriptors were classified as absent (A), normal (N), reduced (R), or increased (In), and NA when descriptors could not be meaningfully evaluated due to the absence of reference waves (Eggermont JJ, 2019).

### Generation of the theoretical ABR configuration space

All theoretical combinations of the nine descriptors were generated by computing the Cartesian product of their possible categorical states. At this stage, descriptors were initially treated as independent variables. This assumption did not imply physiological independence, but was adopted to ensure exhaustive enumeration of the complete theoretical outcome space prior to applying formal constraints. The initial combinatorial expansion thus represented the maximal abstract ABR space from which implausible or redundant configurations could subsequently be removed.

### Internal dependencies and duplicate configurations

Several ABR descriptors are meaningful only when the corresponding waves are identifiable, else the model explicitly incorporates non-applicable values (NA). For instance, if a wave was absent, its latency could not be evaluated. Similarly, amplitude ratios and interpeak intervals involving absent waves could not be interpreted. These dependencies were resolved prior to diagnostic annotation by substituting non-applicable values with a dedicated placeholder.

The following substitutions were applied: if wave I was absent, LI was considered non-applicable; if wave III was absent, LIII was considered non-applicable; if wave V was absent, LV was considered non-applicable; if either wave I or wave V was absent, the I/V ratio and the IT (between I and V) interval were considered non-applicable; and if either wave III or wave V was absent, the IB (between III and V) interval was considered non-applicable.

The presence of non-applicable values enabled the individuation of formally different but physiologically equivalent configurations, preventing them from being treated as distinct diagnostic states, and allowing to prune duplicates.

### Constraints definitions and configuration pruning

The complete theoretical configuration space was progressively reduced through a predefined pruning procedure.

The first level of pruning addressed redundancy. After non-applicable values were assigned, several configurations became functionally identical, particularly when the absence of a wave rendered downstream descriptors uninterpretable. Redundant rows were therefore collapsed, ensuring that each retained configuration represented a unique, non-overlapping state in the ABR diagnostic space.

The second level of pruning addressed algebraic consistency. Configurations were excluded when derived descriptors were mathematically incompatible with their constitutive variables. Specifically, interpeak intervals were required to remain consistent with the corresponding absolute latencies, and the I/V amplitude ratio had to remain consistent with the relative states of waves I and V (e.g., if both I and V waves present normal amplitudes, their ratio will necessarily be normal, as well). This step ensured that the retained configurations adhered to the internal logic of the descriptor system.

The third level of pruning addressed neurobiological plausibility. Configurations implying physiologically impossible conduction patterns were removed. Specifically, decreased latencies were considered incompatible with the known biophysical properties of the auditory pathway, as they would imply conduction velocities exceeding plausible physiological limits or indicate technical error or wave misidentification rather than a meaningful diagnostic condition (Oyler et al., 1991). This filtering step ensured that the diagnostic space was constrained by both formal logic and auditory neurophysiology. The final set of valid configurations constituted the theoretical domain for expert annotation and rule extraction.

The formalization of the described steps in Boolean logic is provided in the Supplementary Material (Supplementary Material 1), alongside the R code implementing it (Supplementary Material 2).

### Diagnostic categories

A set of canonical diagnostic categories was defined to capture the principal clinically interpretable patterns of ABR abnormality. These categories were selected to represent both peripheral and central dysfunctions while maintaining a level of granularity compatible with rule-based interpretation. The diagnostic labels included: Normal (NORM); Pathological Peripheral Severe Hypoacusia/Anacusia (PPSHA); Pathological Peripheral Conductive Hypoacusia (PPC); Pathological Peripheral Sensory-Neural (PPSN); Pathological Peripheral Retro-Ganglionic abnormality (PPRG); Pathological Brainstem abnormality (PB); and Mixed abnormality (MIXED) (Young et al., 2023).

The NORM category represented the unique configuration in which all relevant descriptors were within normal limits. PPSHA captured severe peripheral auditory impairment or anacusis, typically reflected by the absence of the main ABR waves. PPC represented a conductive pattern in which wave morphology and interpeak transmission were preserved, but absolute latency abnormalities indicated delayed sound transmission, as observed in otitis media (McGee and Clemis, 1982). PPSN denoted sensory-neural peripheral dysfunction, a type of hearing loss originating in the inner ear, including the cochlea and associated structures (Liberman and Kujawa, 2017). PPRG indicated retro-ganglionic involvement, encompassing patterns compatible with proximal auditory nerve dysfunction. PB represented abnormalities affecting central auditory conduction through the brainstem (Eggermont JJ, 2019). MIXED was reserved for configurations suggesting more than one mechanism or for patterns not meeting criteria for a more specific category.

### Expert-based annotation of valid configurations

All valid configurations were independently reviewed by two expert raters with experience in clinical neurophysiology and ABR interpretation. Raters were provided with the descriptor values for each configuration and assigned a diagnostic label from the predefined categories. Annotation was performed at the level of theoretical feature combinations rather than waveform images, as the aim of this phase was to formalize diagnostic reasoning from structured neurophysiological descriptors. The classification provided by Rater 1 (DRS) served as the primary reference for rule extraction, while the classification by Rater 2 (DC) was used to assess the consistency of expert reasoning and to support validation of the proposed diagnostic structure. Inter-rater agreement was quantified as percentage agreement; disagreements were considered informative, as they identified regions of the diagnostic space where the theoretical mapping was less intuitive or where competing physiological interpretations were possible.

### Classification rule extraction

Classification rules were extracted to translate expert diagnostic reasoning into an explicit, reproducible decision framework. Two complementary approaches were employed. First, an exploratory procedure based on a LLM was applied to the expert-labelled configurations. The model, specifically GPT-5.5 (OpenAI, 2026), was prompted to infer candidate hierarchical rules capable of reproducing the previously uploaded diagnostic assignments. This step served as a support for pattern discovery and rule formulation, rather than as an autonomous source of ground truth. We refer to this classification as “LLM rules”.

Second, a logic-based iterative procedure was conducted by progressively examining descriptor values and combinations uniquely associated with specific diagnostic labels. Simple and highly specific patterns were identified first, followed by increasingly complex combinations of descriptors. This procedure enabled the construction of a hierarchical rule set in which more specific diagnostic conditions were evaluated before broader residual categories. In the resulting structure, normal and severe peripheral absence patterns were assigned first, followed by specific peripheral conductive, retro-ganglionic, sensory-neural, and brainstem patterns. Configurations not satisfying any of the previous rules were assigned to the MIXED category. We refer to this classification as “Logic rules”.

### Comparison and characterization of extracted rules

The classifications generated by the rule-based procedure were compared with expert annotations to evaluate the extent to which the formal model reproduced expert reasoning. Agreement was summarized using percentage concordance, and the unweighted coefficient Cohen’s kappa, as diagnostic labels were treated as nominal categories. Cohen’s kappa was considered to account for chance agreement between classifying agents (including both raters, and both algorithmic classifications), in a pairwise manner. We retained as final rule set the one presenting the higher average Cohen’s k, with respect to the raters.

The distribution of diagnostic labels across the final valid configuration space was described to characterize the relative size of each diagnostic region, across the classification proposed by the raters, and by the extracted rules (hereby collectively referred to as “classification agents”).

The distribution of descriptors across diagnostic labels, across classification agents, is assessed, as well, to visualize the descriptors leading to discrepancies in label assignment.

To quantify the contribution of individual descriptors to diagnostic assignment, a Random Forest (RF) classifier was fitted using the diagnostic labels as outcomes and the ABR descriptors as predictors, through the R library random forest v 4.7-1.1 (Liaw and Wiener, 2002). This analysis was not intended to replace the rule-based model or to provide an alternative black-box classifier. Instead, it was used descriptively to estimate the relative importance of each descriptor in producing the final classification structure, and to compare their importance across classification agents. Following a standard approach, the variable importance was assessed by evaluating the decrement in the Gini Index and in the Mean Accuracy when excluding the variable under consideration, larger decrements are associated with greater variable contribution, and, hence, higher importance (Cutler et al., 2007). The resulting importance profile supported the interpretation of which ABR features most strongly shaped the diagnostic space.

We propose as final rule system the one presenting the higher average Cohen’s kappa with the raters’ classification; this final set was implemented in R to ensure reproducibility and to allow systematic assignment of all valid configurations. In the Supplementary Material, we include all the labels, as assigned by Rater 1 and 2, and according to LLM and Logic Rules.

### Software and reproducibility

All combinatorial generation, pruning procedures, rule implementation, agreement analyses, and descriptive modeling steps were implemented in R version 4.3.2 (R Core Team, 2023). The code used to generate the theoretical combinations, highlight non-applicable values, remove invalid or redundant configurations, and implement diagnostic rules was provided in Supplementary Material 2.

## RESULTS

### Construction and Refinement of the Theoretical ABR Configuration Space

The combinatorial expansion of nine predefined ABR descriptors produced an initial theoretical space of 5832 possible configurations, representing the maximal set of formal descriptor combinations prior to the application of dependency rules, algebraic constraints, and neurobiological plausibility criteria. Following the assignment of non-applicable values, 4250 configurations were identified as redundant and were discarded, resulting in 1582 unique configurations for further analysis. Of these, 384 were excluded due to violations of algebraic consistency rules between primary and derived descriptors, additional 855 configurations were removed for exhibiting neurobiologically implausible patterns, primarily involving reduced latencies or combinations incompatible with established auditory pathway conduction. The final constrained diagnostic domain comprised 343 valid ABR configurations. Figure 1 illustrates the progressive pruning procedure, and the number of retained and discarded combination at each step.

**Figure 1.**
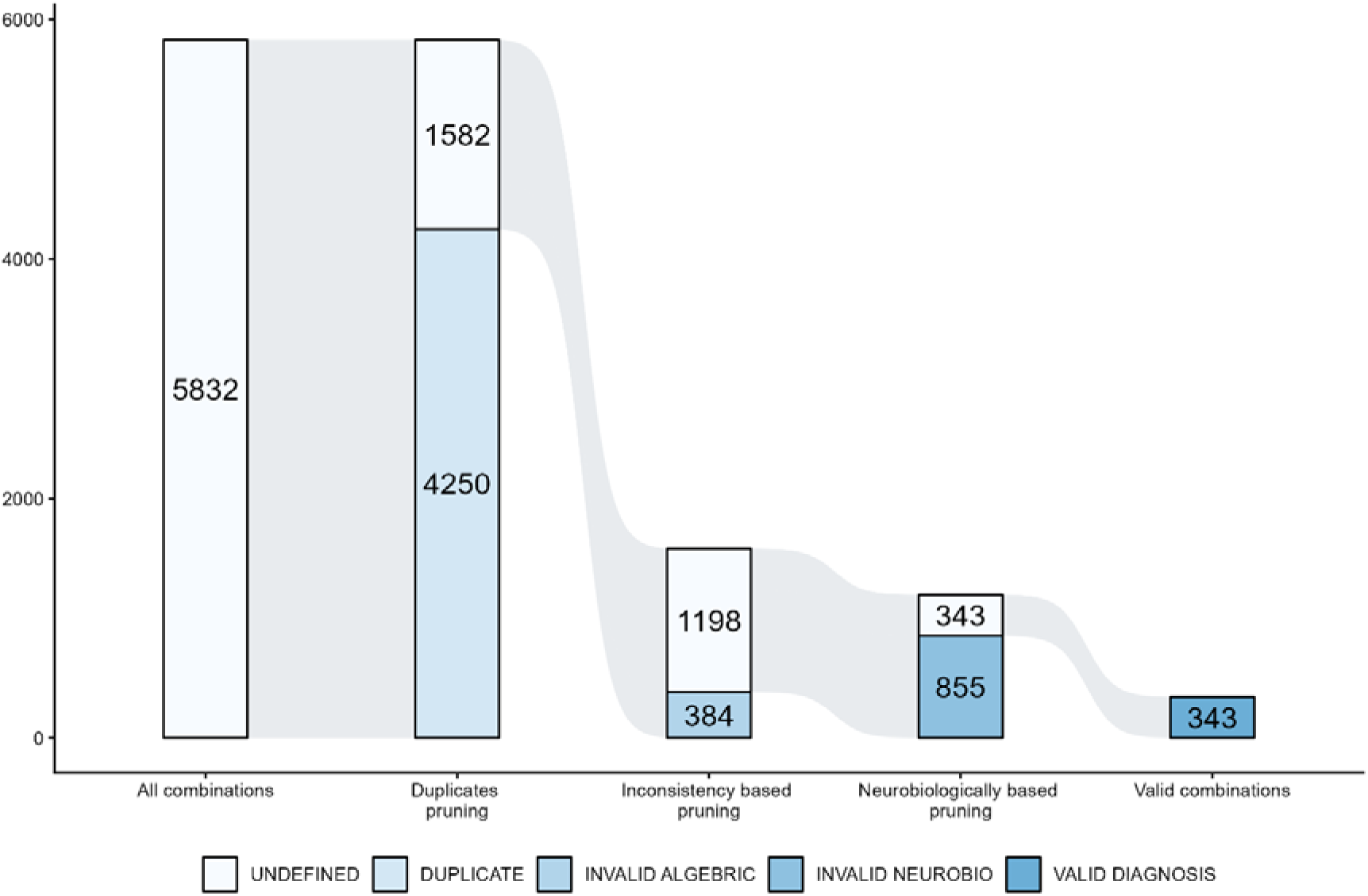
Progressive pruning of combinations.

Supplementary Material 3 contains the complete list of generated configurations, the annotation leading to pruning, and the retained valid configurations.

### Agreement Between Expert Raters and Algorithmic Classification

Agreement analyses were conducted across the 343 valid annotated ABR configurations. The two expert raters showed high concordance: Rater 1 and Rater 2 agreed on 289 of 343 configurations, corresponding to 84.55% agreement and a Cohen’s κ of 0.620. Agreement between expert classifications and the LLM classification was lower: Rater 1 agreed with the LLM on 215 of 343 configurations (62.68% agreement; Cohen’s κ = 0.356), while Rater 2 on 203 of 343 configurations (59.18% agreement; Cohen’s κ = 0.324). The agreement with Logic rules was higher for both Rater 1 (76.97% agreement; Cohen’s κ = 0.533) and Rater 2 (73.18% agreement; Cohen’s κ = 0.490). Finally, the two automatic rule-based system agree on 83.38% of combinations, with Cohen’s κ = 0.727. Table 1 presents the agreement percentage and Cohen’s k across the considered classification systems.

**Table 1.** Agreement percentage and Cohen’s K across the considered classifications systems.

|  | RATER 1 | RATER 2 | LLM RULES |
| --- | --- | --- | --- |
| RATER 2 | 84.55%; $\kappa = 0.620$ | - | |
| LLM RULES | 62.68%; $\kappa = 0.356$ | 59.18%; $\kappa = 0.324$ | - |
| LOGIC RULES | 76.97%; $\kappa = 0.533$ | 73.18%; $\kappa = 0.490$ | 83.38%; $\kappa = 0.727$ |

### Extracted Hierarchical Classification Rules

Overall, since Logic rules present the higher average Cohen’s κ, they represent the final set of proposed rules. The rules were hierarchically ordered from the most specific and physiologically constrained to broader residual classification. A logical formalization of Logic rules, and their R implementation is provided in the Supplementary Material 4 and 2, respectively, and hereby reported in natural language:

1. NORM: assigned when all considered parameters are equal to N.
2. PPSHA: assigned when waves I, III, and V are A.
3. PPC: assigned when waves I, III, and V, the intervals IT, IB and the I/V ratio are all equal to N, while LI is In.
4. PPRG: assigned when waves I, III, and V are classified as N, the I/V ratio is not R, LI and IB are classified as N, and at least one of the following features is classified as In: I/V ratio, LIII, or IT.
5. PPSN: assigned when waves III, V, and LV are classified as N, the I/V ratio is not In, IB is classified as N, and IT is not classified as In.
6. PB: assigned when wave I is classified as N, the I/V ratio is not R, and either:

a. wave III is classified as A, or
b. wave III is classified as N with LI classified as either N or In.
7. MIXED: assigned when none of the above conditions are met.

The NORM rule identified the unique configuration in which all descriptors were normal. The PPSHA rule identified configurations in which waves I, III, and V were absent, regardless of the remaining non-applicable descriptors. The PPC rule identified conductive patterns characterized by preserved wave amplitudes and preserved interpeak conduction, with increased absolute latency of wave I. The PPRG rule captured configurations with preserved waves I, III, and V, normal wave I latency, and normal III-V interval, but with abnormalities in at least one descriptor reflecting altered relationships between early and later components. The PPSN rule identified peripheral sensory-neural patterns characterized by preserved waves III and V, preserved wave V latency and III-V interval, and absence of criteria indicating increased total brainstem conduction time.

Two complementary PB rules were required to capture brainstem patterns. The first identified configurations with preserved wave I, absent wave III, and an I/V amplitude ratio not consistent with a reduced peripheral pattern. The second identified configurations with preserved waves I and III, an I/V ratio not reduced, and normal or increased wave I latency, following exclusion of NORM, PPC, and PPRG categories. All configurations not satisfying any of the preceding criteria were assigned to the MIXED category.

Supplementary Material 5 provides the complete list of valid configurations and the diagnostic labels assigned by both raters and both rules systems.

### Distribution of Diagnostic Labels across Raters and Rules Systems

The distribution of assigned diagnostic labels varied between the two expert raters and between the two proposed rule-based classification approaches (see Figure 2). In both expert and algorithmic classifications, NORM, PPC, and PPSHA occupied small, narrowly defined regions, while MIXED and PB constituted the largest regions of the diagnostic space.

**Figure 2.**
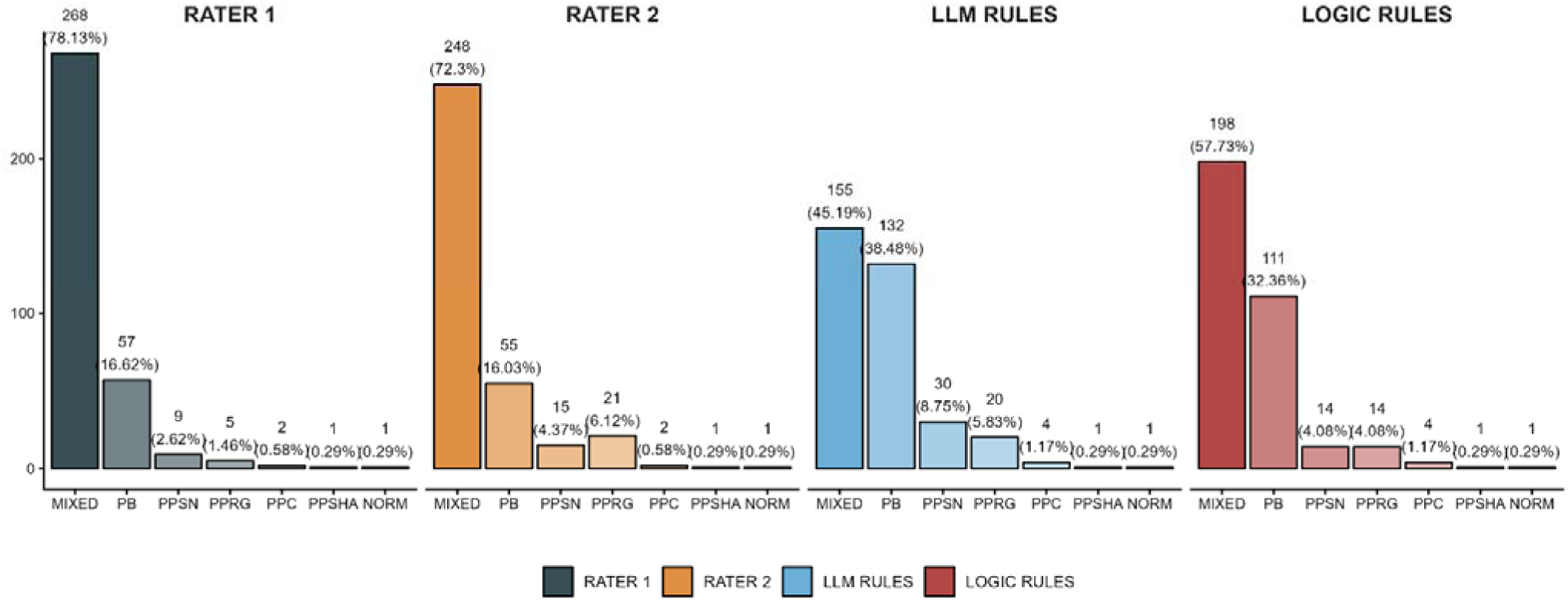
Distribution of diagnostic Labels across raters and classification approaches. Legend: LLM = Large Language Model; MIXED = mixed abnormalities; PB = brainstem abnormalities; PPSN = sensori-neural hypoacusia; PPRG = retro-ganglionic abnormalities; PPC = conductive hypoacusia; PPSHA = severe hypoacusia/anacusia; NORM = normal.

The two expert raters exhibited a similar overall distribution, with a predominance of MIXED and PB labels. This is true, as well, for both the proposed sets of rules. Nonetheless, the prevalence of MIXED labels is less predominant, in favour of a higher proportion of combinations labelled as PB.

### Descriptor Profiles Associated with Diagnostic Labels

Figure 3 presents the distribution of descriptor values across diagnostic labels, as proposed by raters and rule-based systems.

**Figure 3.**
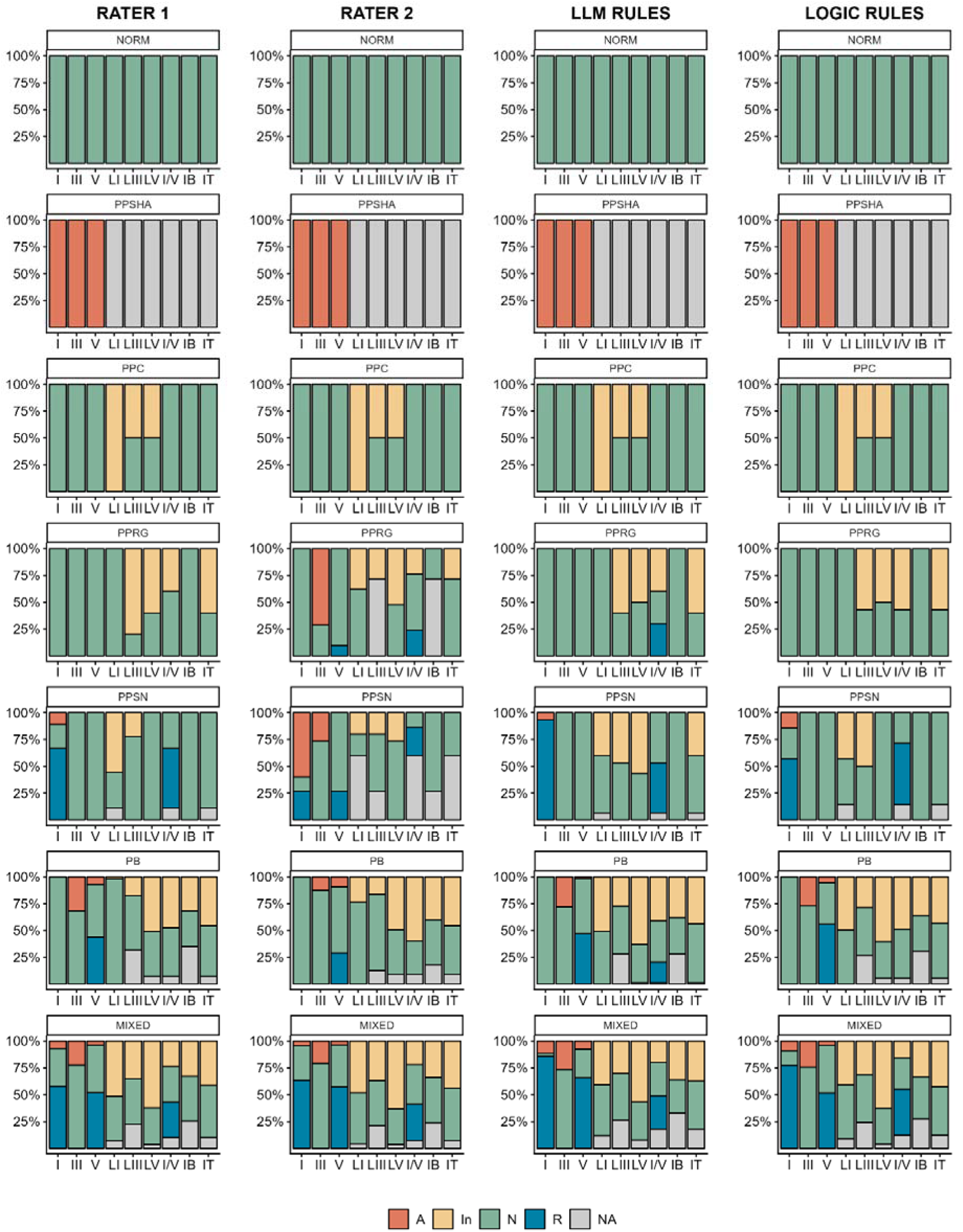
The distribution of descriptor values across diagnostic labels. Legend: LLM = Large Language Model; MIXED = mixed abnormalities; PB = brainstem abnormalities; PPSN = sensori-neural hypoacusia; PPRG = retro-ganglionic abnormalities; PPC = conductive hypoacusia; PPSHA = severe hypoacusia/anacusia; NORM = normal. I = amplitude of I component; III = amplitude of III component; V = amplitude of V component; I/V amplitude ratio of I and V components; LI = latency of I component; LIII = latency of III component; LV = latency of V component; IB = brainstem interval (difference between LIII and LV); IT = total interval (difference between LI and LV). A = absent; In = increased; N = normal; R = reduced; *** = not applicable.

The NORM category corresponded to a single homogeneous configuration in which all amplitudes, latencies, amplitude ratio, and interpeak intervals were within normal limits. PPSHA was similarly highly constrained and was characterized by the absence of the main ABR waves. In this category, downstream latency and interval descriptors were non-applicable.

The PPC category was also narrowly defined, as well, and consistently characterized across agents by preserved wave amplitudes and preserved interpeak conduction, combined with increased absolute latencies. This pattern was consistent with a conductive mechanism in which peripheral sound transmission is delayed while brainstem conduction remains preserved.

The PPRG category included configurations with preserved wave presence but abnormalities in the relationship between early and later ABR components, particularly the I/V amplitude ratio and latency-derived descriptors. This pattern was consistent with retro-ganglionic involvement, in which proximal auditory nerve or early post-synaptic dysfunction may alter the relative contribution of wave I and later brainstem components.

The PPSN category was primarily associated patterns in which later waves and central conduction descriptors could remain preserved despite abnormalities in early peripheral components due to the loss of earing cells.

The PB category encompassed a broader and more heterogeneous region, including configurations compatible with altered central auditory conduction through the brainstem, particularly when abnormalities involved wave III, the I/V amplitude ratio, or interpeak relationships not explained by purely peripheral patterns. The MIXED category included configurations that did not satisfy the specific criteria for the other diagnostic classes, reflecting the coexistence of multiple abnormal features or insufficient specificity for a single interpretation.

### Descriptor Importance Analysis

Random Forest analysis, combined with variable importance extraction, was conducted to identify which ABR features most strongly contributed to diagnostic assignment. Across expert and rule-based classifications, wave I amplitude consistently demonstrated the highest importance, as measured by both mean decrease in Gini index and mean decrease in accuracy.

Other descriptors exhibited different variable importance values depending on the classification agents. For instance, LI results relevant in the classification proposed by Rater 1, while V amplitude and IB are more important for the LLM based rules. Figure 4 presents the variables importance for the diagnostic labels of the two raters and the rules based systems.

**Figure 4.**
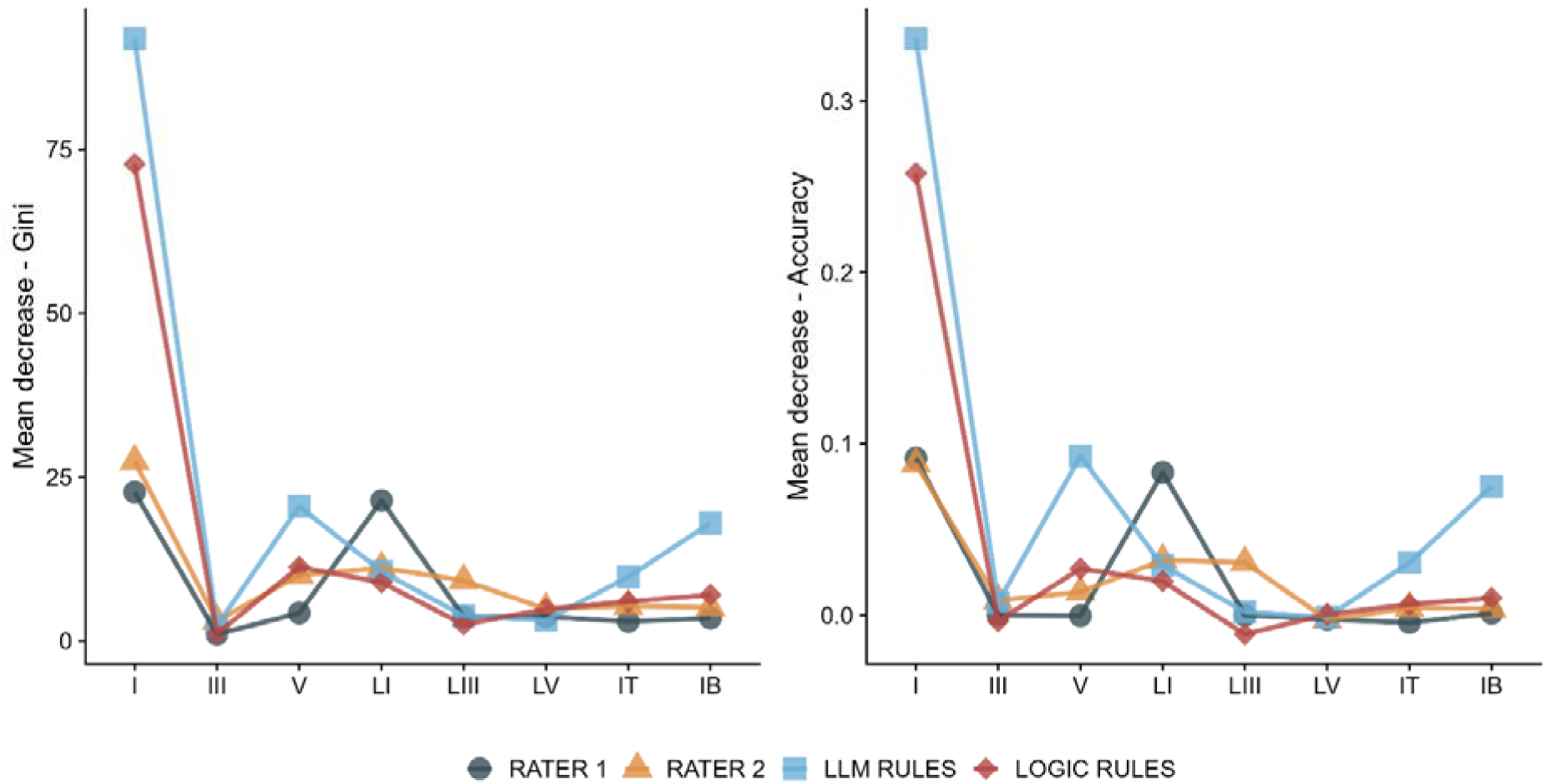
Variable importance for the labelling, across classification agents. Legend: LLM = Large Language Model; I = amplitude of I component; III = amplitude of III component; V = amplitude of V component; I/V amplitude ratio of I and V components; LI = latency of I component; LIII = latency of III component; LV = latency of V component; IB = brainstem interval (difference between LIII and LV); IT = total interval (difference between LI and LV).

## DISCUSSION

In this study, we developed and evaluated a theoretical, rule-based framework for ABR classification. Our principal finding is the reduction of an initial combinatorial space of 5,832 possible configurations to a final constrained diagnostic domain of 343 valid configurations, achieved through the sequential application of internal dependency rules, algebraic consistency criteria, and neurobiological plausibility constraints. (Wimalarathna et al., 2022) This outcome supports the central premise of the study: ABR interpretation can be formalized as a structured diagnostic space rather than being viewed solely as a collection of independent waveform abnormalities.

The proposed framework targets a level of ABR automation that differs from most previous computational approaches. Prior studies have demonstrated that ABR analysis can be automated for threshold detection, response identification, waveform recognition, and signal feature extraction (Bogaerts et al., 2009; Molina et al., 2016; Suthakar and Liberman, 2019; Chen et al., 2021; McKearney and MacKinnon, 2019; McKearney et al., 2022; Thalmeier et al., 2022; Liang et al., 2024; Erra et al., 2026). However, these methods primarily address the detection or quantification of ABR components. In contrast, our work focuses on the subsequent diagnostic step: transforming structured neurophysiological descriptors into clinically interpretable categories.

This distinction is significant because ABR interpretation extends beyond simply determining the presence of waves I, III, or V or identifying abnormal latency or amplitude. Each descriptor gains diagnostic significance only in relation to the others, mirroring routine clinical reasoning. Absolute latencies, interpeak intervals, wave amplitudes, and amplitude ratios must be interpreted collectively to assess whether the observed pattern aligns with a functional and anatomical classification (Young et al., 2023). Our framework seeks to make this reasoning explicit, reproducible, and computationally implementable.

### Interpretation of the constrained diagnostic space

The pruning procedure constitutes a primary conceptual contribution of this study. Beginning with the full Cartesian product of the nine ABR descriptors, we eliminated a substantial number of configurations that were redundant, algebraically inconsistent, or neurobiologically implausible. This process demonstrates that the theoretical ABR space is not an unconstrained multidimensional domain, but is instead strongly shaped by internal dependencies among wave presence, latencies, amplitude ratios, and interpeak intervals (Molina et al., 2016).

Redundant configurations were excluded primarily because several descriptors become non-applicable when the corresponding waves are absent. For instance, if wave I or wave V is absent, the I/V ratio and the I-V interval cannot be physiologically interpreted. Similarly, the absence of wave III or wave V precludes the III-V interval from serving as an independent diagnostic feature. By introducing inapplicable values, we were able to collapse formally distinct but physiologically equivalent configurations into a single diagnostic state.

Eliminating algebraically inconsistent configurations further enhanced the model’s internal coherence. Derived descriptors, such as amplitude ratios and interpeak intervals, cannot vary independently from their underlying variables. For example, the I/V amplitude ratio must be compatible with the relative states of waves I and V, and interpeak intervals must align with the corresponding absolute latencies (Young et al., 2023). This step is essential for transforming ABR interpretation into a reproducible, rule-based system.

Finally, neurobiological pruning eliminated configurations incompatible with established auditory pathway conduction. Reduced latencies were deemed implausible as meaningful diagnostic abnormalities, as they would suggest conduction velocities beyond physiological limits or, more likely, technical error or wave misidentification (Oyler et al., 1991). Thus, the final set of 343 valid configurations reflects both formal logic and auditory neurobiology (Eggermont, 2019; Young et al., 2023).

### Diagnostic asymmetry and the meaning of the MIXED category

The final diagnostic space exhibited marked asymmetry. NORM, PPSHA, and PPC occupied narrow, highly constrained regions, while PB and MIXED encompassed much larger portions of the diagnostic domain. This distribution is clinically plausible. A normal ABR is defined by a highly specific configuration in which all relevant descriptors fall within normal limits. Likewise, severe hypoacusia or anacusia is represented by a stereotyped absence pattern, and conductive hypoacusia is characterized by a specific combination of preserved waves and interpeak conduction with delayed absolute latency (McGee and Clemis, 1982).

In contrast, brainstem and mixed abnormalities are combinatorially broader, resulting from various combinations of altered wave morphology, delayed conduction, abnormal interpeak intervals, or altered relationships between early and later components. Mixed abnormalities are even more expansive, as they encompass configurations where multiple mechanisms may coexist or where available descriptors are insufficient to distinguish among them, resulting in differing interpretations of the data.

This result underscores a key principle of diagnostic neurophysiology: normality occupies a narrow domain, while abnormality is combinatorically broad (Krumbholz et al., 2020). An isolated descriptor may have different implications depending on the broader configuration in which it appears. For instance, increased absolute latencies with preserved interpeak intervals may indicate a conductive mechanism, whereas abnormalities involving interpeak relationships may suggest retro-ganglionic or brainstem dysfunction. Our framework captures this contextual aspect of ABR interpretation by assigning diagnostic labels to complete configurations rather than to isolated features.

The large size of the MIXED category should not be viewed solely as a limitation. It also reflects a realistic aspect of expert interpretation. In clinical practice, certain ABR patterns cannot be confidently attributed to a single anatomical site or pathophysiological mechanism. Mixed configurations may indicate combined peripheral and central involvement, partial wave absence, ambiguous latency relationships, or patterns where multiple diagnostic hypotheses remain plausible. By explicitly defining a MIXED region, our framework preserves diagnostic caution rather than forcing all configurations into overly rigid categories.

### Expert agreement and rule-based classification

The two expert raters demonstrated high agreement across the final set of valid configurations, with 84.55% concordance and Cohen’s κ = 0.620 (Naves et al., 2012). This finding supports the clinical interpretability of the constrained diagnostic space. The raters classified abstract combinations of structured descriptors rather than raw ABR traces. Their agreement indicates that a substantial portion of expert ABR reasoning can be reproduced at the level of formalized neurophysiological features.

Agreement between expert classifications and LLM rules was lower, with Cohen’s κ = 0.356 for Rater 1 and κ = 0.324 for Rater 2 (Naves et al., 2012). This suggests that the LLM-derived candidate rules captured only a portion of expert reasoning. This result is informative, as it demonstrates that language-model-assisted extraction can identify plausible classification structures, but should not be regarded as a sufficient source of clinical ground truth (Jedrzejczak et al., 2026). In this study, the LLM served as an exploratory tool for rule discovery rather than as an autonomous diagnostic classifier.

The Logic rules demonstrated higher agreement with both raters than the LLM rules, with Cohen’s κ = 0.533 for Rater 1 and κ = 0.490 for Rater 2. Additionally, Logic rules and LLM rules agreed in 83.38% of configurations, with Cohen’s κ = 0.727 (Naves et al., 2012). These findings indicate that both automatic procedures captured a partially shared structure of the diagnostic space, but the final logic-based rule system more closely approximated expert classification. Consequently, the Logic rules were retained as the final proposed rule set.

Discrepancies between expert raters and automatic rules are pivotal, as they highlight areas of the diagnostic space where the current formalization is not fully conclusive and where data interpretation remains debatable. The most challenging boundary involves PB and MIXED configurations. This is clinically plausible, since brainstem conduction abnormalities often require interpretation of multiple interdependent features, and distinguishing them from mixed peripheral-central patterns may not always be reducible to deterministic rules. These regions should be prioritized in future refinement efforts.

### Relationship with previous automated ABR approaches

The present study should be interpreted as complementary to previous work on automated ABR analysis. Automated threshold detection methods have aimed to reduce subjectivity and variability in visual threshold estimation (Bogaerts et al., 2009; Suthakar and Liberman, 2019; Tanaka et al., 2023; Thalmeier et al., 2022). Machine-learning and deep-learning models have shown promising results in response classification, detection of characteristic waveforms, and automated recognition of ABR components (McKearney and MacKinnon, 2019; Chen et al., 2021; McKearney et al., 2022; Liang et al., 2024). More recent approaches have extended automation toward hearing-loss classification and open-source deep-learning pipelines for peak amplitude, latency, and threshold extraction (Ma et al., 2024; Erra et al., 2026).

Our work diverges from these approaches by not primarily addressing signal detection. Instead, we formalize the diagnostic implications of already-defined ABR descriptors. In a potential future pipeline, automated waveform analysis tools could extract waves, amplitudes, latencies, ratios, and intervals from raw recordings. These outputs could then be processed by our diagnostic layer, which would provide an interpretable classification and a transparent rule path to the final label.

This downstream role is clinically significant. A model may accurately detect wave V, estimate thresholds, or classify the presence of a response, yet still fail to explain why a particular combination of wave abnormalities should be interpreted as conductive, sensory-neural, retro-ganglionic, brainstem, or mixed. The primary contribution of our model lies in this explanatory layer. It transforms ABR automation from a purely signal-processing task into a diagnostic reasoning framework.

### Interpretability and symbolic value

Interpretability is a principal strength of the proposed model. In clinical neurophysiology, black-box outputs are generally insufficient, even when statistical performance is high. Clinicians require an understanding of why a specific diagnostic label has been assigned, which descriptors contributed to the decision, and whether the classification aligns with established pathophysiological processes (Norrix and Velenovsky, 2018). This requirement is especially important for evoked potentials, where diagnostic meaning depends on the relationships among multiple descriptors rather than on a single measurement.

The proposed approach aligns with ongoing discussions on explainable artificial intelligence in medicine. Clinical decision-support systems are increasingly expected to be not only accurate, but also transparent, reproducible, and compatible with clinical workflows (Combi et al., 2022; Susanto et al., 2023; Elhaddad and Hamam, 2024).

Our work can thus be regarded as a symbolic framework, as it integrates computational formalization with explicit neurophysiological knowledge. The diagnostic categories are generated not by uninterpretable statistical associations alone, but by rules that can be inspected, discussed, modified, and clinically evaluated. This is particularly relevant for future implementation, since a decision-support system in neurophysiology should provide not only an answer but also the reasoning underlying it.

### Role of individual ABR descriptors

The importance of individual descriptors was highlighted by the RF analysis, which offered a complementary characterization of the diagnostic space. In an interpretable classification framework, it is essential not only to assign a diagnostic label but also to identify which variables most strongly contribute to that assignment. This information is clinically relevant, as it clarifies whether the diagnostic decision is primarily influenced by early peripheral descriptors, later brainstem components, latency abnormalities, amplitude relationships, or interpeak conduction measures. Descriptor-importance analysis thus provides a quantitative estimate of which neurophysiological features carry the greatest diagnostic weight within the proposed theoretical space.

Across both expert and rule-based classifications, wave I amplitude consistently demonstrated the highest importance, as indicated by both the mean decrease in the Gini index and the mean decrease in accuracy (Kamerer et al., 2020). This finding is physiologically meaningful. Wave I represents the earliest ABR component and reflects distal auditory nerve activity. Alterations in wave I therefore have a strong impact on distinguishing between peripheral and non-peripheral patterns (Young et al., 2023). The significance of wave I aligns with the role of early auditory nerve activity in peripheral auditory dysfunction and the importance of cochlear and synaptic mechanisms in sensory-neural hearing loss (Liberman and Kujawa, 2017).

More broadly, these findings indicate that even a limited number of well-selected ABR descriptors can suffice to identify clinically meaningful diagnostic patterns. The model does not require the full complexity of the waveform to extract useful diagnostic information. Instead, it depends on a small set of highly informative variables whose combinations constrain the range of plausible interpretations. For example, recognizing a feline from a limited anatomical feature, such as the tail, may sometimes be sufficient to distinguish a lion from a tiger; when this distinction is not possible, the same feature may still help exclude other possibilities, such as a lynx. Similarly, a restricted set of ABR descriptors may not always enable the model to assign the most specific diagnosis, but it can still narrow the diagnostic field by excluding physiologically incompatible patterns.

Other descriptors exhibited varying importance depending on the classification agent. Specifically, LI contributed significantly to the classification proposed by Rater 1, while wave V amplitude and IB were more influential in the LLM-based rules. This variability suggests that different classification agents may employ distinct diagnostic strategies, even within the same constrained descriptor space. Rather than undermining the framework, this finding helps identify which features drive convergence or divergence between expert and algorithmic interpretations.

Simultaneously, the contribution of interpeak intervals and later components underscores the central role of conduction measures in identifying retro-ganglionic and brainstem abnormalities. The I-V and III-V intervals are not merely isolated numerical values, but descriptors of functional conduction across successive segments of the auditory pathway. Their diagnostic significance depends on the states of the corresponding waves and their relationships to absolute latencies and amplitude ratios (Eggermont, 2019; Young et al., 2023). Overall, the descriptor-importance analysis supports the physiological plausibility of the proposed diagnostic architecture and confirms that our model is guided by clinically interpretable variables rather than opaque statistical associations.

### Future outlook

The most critical future step is validation using real-world ABR recordings. In such a pipeline, raw ABR traces could first be processed by automated or semi-automated tools for wave detection, latency estimation, amplitude measurement, and threshold identification. The extracted descriptors could then be input into our diagnostic layer, which would assign a diagnostic category and provide the corresponding rule path. This approach would integrate data-driven signal analysis with interpretable neuro-symbolic reasoning.

A second future direction involves reducing diagnostic uncertainty. Instead of generating only a single diagnostic label, future versions of the framework could provide a ranked set of possible interpretations or a confidence score, particularly for configurations near the boundary between PB and MIXED, or between peripheral and retro-ganglionic abnormalities. This approach would more closely mirror clinical reasoning, where some ABR patterns require cautious interpretation and integration with audiological and radiological findings.

A third area for development is adapting the model to clinical context. ABR latencies and thresholds vary with age, maturation, stimulus parameters, acquisition settings, and laboratory normative ranges (Chhajed et al., 2022; Young et al., 2023). Future versions should therefore incorporate age-specific and protocol-specific normative values, enhancing the framework’s applicability to neonatal screening, pediatric assessment, adult neurophysiology, and experimental settings.

### Educational role and learning on the job

The model may also serve an educational purpose. Because each diagnostic label is linked to explicit descriptors and hierarchical rules, our model could be used to train residents, technicians, and junior neurophysiologists in ABR interpretation. It can help learners understand how individual abnormalities acquire meaning only within a broader neurophysiological pattern. In this way, the framework may function not only as a decision-support tool, but also as a structured representation of expert knowledge.

The main contribution of this work is both theoretical and operational. It provides a formal structure that can be implemented, tested, refined, and eventually integrated with automated ABR signal-processing tools. The future value of this framework will depend on its ability to improve reproducibility, support expert interpretation, and reduce variability in clinical ABR reporting.

### Limitations

This study has several limitations. First, the framework was developed and tested using theoretical ABR configurations rather than real-world clinical recordings. As a result, the current findings demonstrate internal coherence, interpretability, and reproducibility of the diagnostic space, but do not yet establish clinical diagnostic accuracy. Clinical ABR recordings introduce additional sources of variability, such as noise, patient age, hearing threshold, stimulus parameters, electrode montage, waveform reproducibility, technical quality, and laboratory-specific acquisition procedures.

Second, the descriptor set was intentionally limited to nine clinically meaningful variables. This decision facilitated interpretability and enabled exhaustive modeling of the diagnostic space, but may have excluded relevant information. Future versions of the framework could incorporate waveform morphology, absolute amplitude values, side-to-side asymmetry, intensity-dependent changes, test-retest reproducibility, and age-adjusted normative values.

Third, the diagnostic categories were designed to represent canonical ABR patterns, but they inevitably simplify clinical reality. The boundaries between sensory-neural, retro-ganglionic, brainstem, and mixed abnormalities may be challenging to define using structured descriptors alone. This limitation is reflected in the size of the MIXED region and the lower agreement between expert classifications and automatic rules.

Fourth, although expert agreement was substantial, the expert annotation relied on only two raters. Additional raters and multicenter expert panels will be necessary to determine whether the proposed diagnostic mapping is generalizable across different neurophysiology laboratories and reporting traditions.

Finally, the language-model-assisted rules should be regarded as exploratory. The LLM was valuable for candidate rule extraction, but was not used as an independent source of diagnostic truth. The final proposed classification system is therefore based on Logic rules, which demonstrated higher average agreement with the raters and can be explicitly implemented and inspected.

## Conclusions

Our work presents an interpretable framework for ABR classification by transforming a broad theoretical combinatorial space into a constrained diagnostic domain. The framework formalizes the relationship between clinically meaningful ABR descriptors and diagnostic categories, identifies regions of agreement and uncertainty between experts and rule-based classifications, and provides a reproducible foundation for future automation. Although validation on clinical ABR recordings is still required, this proof-of-concept study supports the feasibility of modeling ABR interpretation as a structured diagnostic space grounded in auditory neurobiology.

## Data Availability

All data produced are available online at https://github.com/LetiziaClementi/abr_classification_framework

## SUPPLEMENTARY MATERIALS

Supplementary materials are available online at https://github.com/LetiziaClementi/abr classification framework

## Author contributions

Author contributions were defined according to the CRediT (Contributor Roles Taxonomy) framework (Allen et al., 2014).

*Letizia Clementi:* Computation; Conceptualization (supporting); Formal Analysis (leading); Methodology (leading); Software; Visualization (leading); Writing – original draft (supporting). *Daniele Cazzato:* Conceptualization (supporting); Investigation (equal); Writing – review & editing (leading). *Elisa Visani:* Conceptualization (supporting); Formal Analysis (supporting). *Marie Carolina Nicolis di Robilant*: Writing – review & editing (supporting); Visualization (supporting). *Annamaria Gallone:* Writing – review & editing (supporting); Project Administration (supporting). *Virginia Iacobelli*: Writing – review & editing (supporting). *Paola Lanteri:* Funding Acquisition. *Davide Rossi Sebastiano:* Conceptualization (leading); Investigation (equal); Methodology (leading); Project Administration (leading); Supervision; Visualization (supporting); Writing – original draft (leading).

## Acknowledgements

We want to thank the Italian Ministry of Health for supporting our study.

---

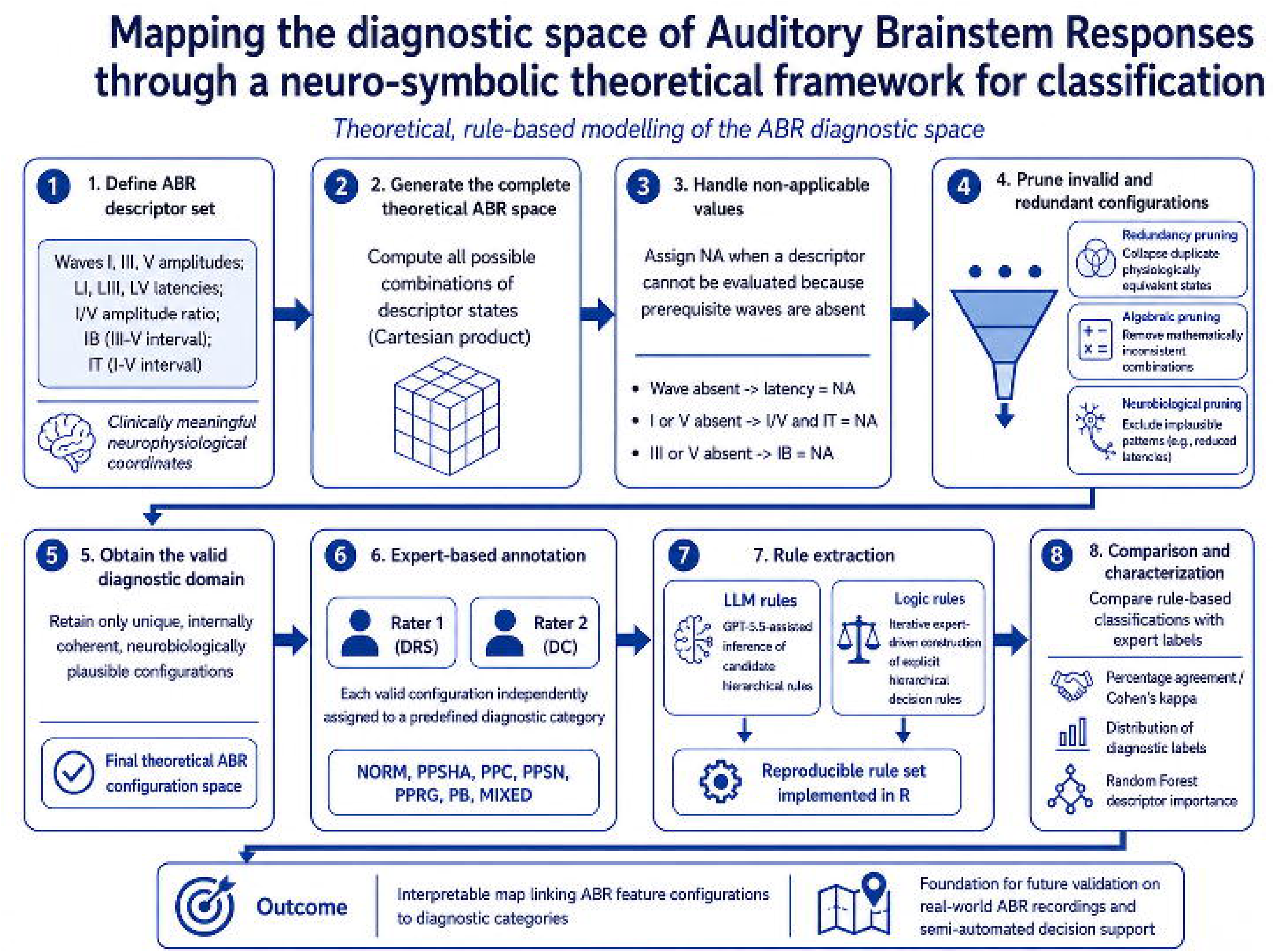

